# Impact of AI-Assisted Mammography Reading on Quality Indicators in the Czech Breast Cancer Screening Programme: A Retrospective Study

**DOI:** 10.64898/2026.05.25.26353869

**Authors:** Lucia Veverková, Zuzana Doležalová, Veronika Maráčková, Eva Mathew, Markéta Urbánková, Monika Ambrožová, Tomáš Piskovský, Ondřej Ngo, Ondřej Májek

## Abstract

**Objectives:** The aim of mammographic screening is the early detection of invasive cancers. In the era of artificial intelligence (AI), this tool may improve diagnosis of earlier stages. The purpose of this study was to assess the impact on selected quality indicators retrospectively.

**Method:** The data source was the Breast Cancer Screening Registry using data from one Screening Unit that currently uses AI routinely. The indicators of the cancer detection rate (CDR), further assessment rate (FAR), and recall rate (RR) in the year 2023, when AI was used, and the year 2022, without AI, in women aged 45-69 were compared. The statistical evaluation used the chi-square test and logistic regression adjusting for the effects of age, a woman’s risk level, and the screening round at a 5% significance level.

**Results:** In 2022, without AI, 4,034 women aged 45–69 were included, compared with 4,049 women in 2023 when AI was used. This study showed a non-significant increase in CDR from 5.0 breast cancers detected per 1,000 women (non-AI assessment) to 5.2 (AI-assisted assessment), *p* = 0.919; OR (95% CI): 1.034 (0.542–1.974), a significant decrease in the FAR from 5.2% to 3.9%, *p* < 0.001; OR (95% CI): 0.665 (0.529–0.836), and a decrease in RR from 2.4% to 1.9%, *p* = 0.083; OR (95% CI): 0.754 (0.548–1.037).

**Conclusion:** AI has the potential to be a useful tool in the early detection of breast cancer by improving quality through a decrease in FAR and RR, while probably maintaining CDR.

**Key points and Clinical Relevance Statement:** *Question:* What is the impact of AI-assissted reading of screening mammography on quality indicators in breast cancer screening programme?

*Findings:* Results of this study shows that using of AI-assisted reading of screening mammography in real clinical practice contributes to reducing further assessment rate and recall rate, while cancer detection rate remains most likely unchanged.

*Clinical Relevance Statement:* AI-assissted reading is an advanced option how to improve the sensitivity and specificity of screening mammography in real-world clinical practice.

## Introduction

The annual incidence of breast cancer in the Czech Republic is approximately 7,700 cases. Although the incidence has increased by 20% (age-standardised European rates, ASR-E), there is a growing proportion of cases diagnosed at earlier stages. In recent years, more than 50% of all breast cancers have been diagnosed at stage 1, and 33% at stage 2; among cancers detected through screening, the proportion of stage 1 breast cancers is even higher, exceeding 75% [1].

The screening programme in the Czech Republic is based on mammography once every two years in women aged over 45 at one of the certified centres [2,3]. Double reading by two radiologists with more than ten years of experience is the standard of care [4]. The performance indicators, such as the recall rate (RR), further assessment rate (FAR), and cancer detection rate (CDR), are evaluated annually. These indicators of the quality are important because an increase in CDR leads to greater sensitivity and a decrease in FAR and RR lead to lower false positivity and to better specificity.

The advances in early radiological detection capabilities and the application of personalised treatment strategies are leading to improved clinical outcomes. The era of artificial intelligence (AI) promises an unprecedented breakthrough in automated imaging analysis methods. As the upcoming future technology, powerful AI systems may be effective at helping radiologists to improve the reduction of false negative and false positive assessments. This has the potential to replace one or both radiologists in the evaluation of screening mammography in the future [5]. These systems are able to identify lesions, distortions or microcalcifications with automatic calculation of the likelihood of the occurrence of breast cancer. Concerns about using AI systems in routine practice stem from their inability to compare mammograms with previous images [6]. AI systems identify some stationary lesions as high-risk. It could lead to increase in number of supplemental examinations. Even when algorithms exhibit high performance in selected research datasets, AI errors in cancer detection such as false positives or false negatives may be greater when algorithms are applied in “real-world” settings or transferred into population-based screening programmes [7].

AI-based software for evaluation of mammography in real clinical practice is used at our center since 2023. This retrospective study shows impact of AI-assissted reading on selected qualitative indicators, especially cancer detection rate, recall rate and further assessment rate, in real clinical practice.

## Methods

This retrospective study was approved by the institutional Ethics Committee of the University Hospital and Faculty of Medicine and Dentistry of Palacký University Olomouc (reference no. 52/26). All procedures were performed in accordance with relevant guidelines and regulations. The patients provided written informed consent to the examination, including consent to using documentation anonymously for scientific and statistical purposes.

This study involves retrospective analyses of one centre in the Czech Republic.

The data source was the Breast Cancer Screening Registry (Institute of Biostatistics and Analyses, Faculty of Medicine, Masaryk University, Brno, Czech Republic) using anonymized data from the Screening Unit of the University Hospital Olomouc. The University Hospital Olomouc has been using AI as an aid or decision support for the standard double reading of mammography images since the beginning of 2023. The aim of the analysis was to compare key quality indicators showing the performance of the programme when using AI vs. without AI.

The AI-based software iCAD SW version 3 was used in real clinical practice on the Senographe Pristina Mammography System (GE HealthCare, 2022).

The inclusion criteria for breast cancer screening programme in Czech Republic are the women being over 45 years old and without symptoms of breast disease. The exclusion criteria are the women being younger than 45, the presence of symptoms of breast disease and mammography in last two years. The analysis considered all the women aged between 45 and 69 years old who underwent screening mammography at the University Hospital Olomouc in 2022 and 2023.

In 2022, mammograms were evaluated with standard double reading by radiologists and without AI-assisted reading, whereas in 2023 they were evaluated with standard double reading by radiologists and already also AI-assisted reading.

The quality indicators, including CDR (the number of invasive and in situ breast cancers, ICD-10 codes C50 and D05, detected per 1,000 women screened), FAR (the proportion of follow-up examinations on days 0–190 after screening mammography), and RR (the proportion of follow-up examinations on days 1–190 after screening mammography, i.e. the woman physically returning to the screening centre for additional examination), were compared between the two time periods.

The following statistical methods (tested at a 5% significance level) were used to compare the indicators in 2022 (without AI assessment) vs. 2023 (with AI assessment): chi-square test and logistic regression, which examined the effect of AI adjusted for age, risk level, and the screening round (initial vs. subsequent) of the women. The women’s risk level is defined as the risk of developing breast cancer and is divided into three categories: women with no increased risk, women with increased risk, and women with no reported risk.

The statistical analysis was performed with the Stata/IC 15.1 software.

## Results

In total, 5,807 images of women without symptoms of breast disease aged 45 years and older were evaluated in 2022 and 5,932 cases in the year 2023 with AI-assisted reading (Figure 1).

**Figure 1:**
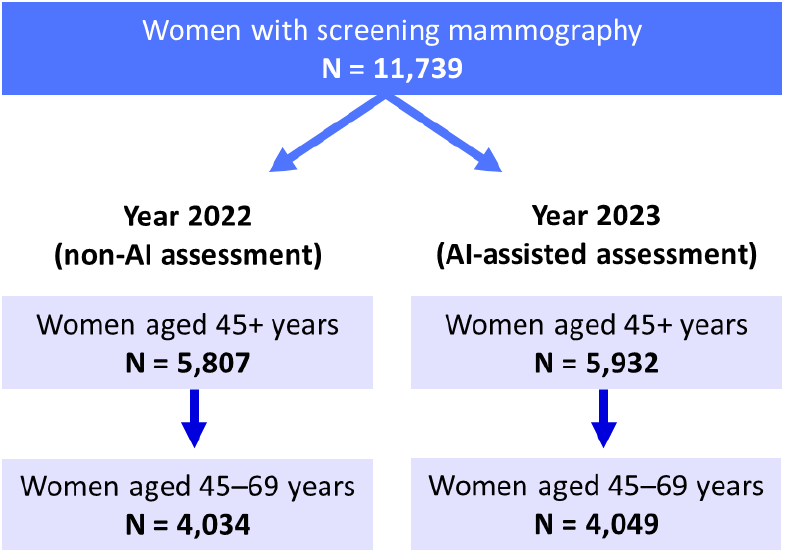
Cohort in 2022 and 2023 due to AI-assistance reading and according to the age of women.

In 2022, 4,034 mammograms were evaluated in women aged between 45 and 69 years old (Table 1). Of this number, 513 women were examined for the first time and 3,521 women were examined in subsequent rounds. There were 3,876 women with no increased risk, 84 women with increased risk, and 74 women with no reported risk. The number of cancers detected was 20. The proportion of women invited for additional examinations in the interval 0–190 days after screening mammography (the further assessment rate) was 5.2% (210 women) and the proportion of women invited for additional examinations in the interval 1–190 days after screening mammography (the recall rate) was 2.4% (98 women) in the evaluation year 2022. In 2023, 4,049 mammograms were evaluated in women aged between 45 and 69 years old (Table 1). Of this number, 539 women were examined for the first time and 3,510 women were examined in subsequent rounds. There were 3,511 women with no increased risk, 73 women with increased risk, and 465 women with no reported risk. The number of cancers detected in the evaluation cohort in 2023 was 21. The proportion of women invited for additional examinations in the interval 0–190 days after screening mammography (the further assessment rate) was 3.9% (157 women) and the proportion of women invited for additional examinations in the interval 1–190 days after screening mammography (recall rate) was 1.9% (75 women).

**Table 1:**
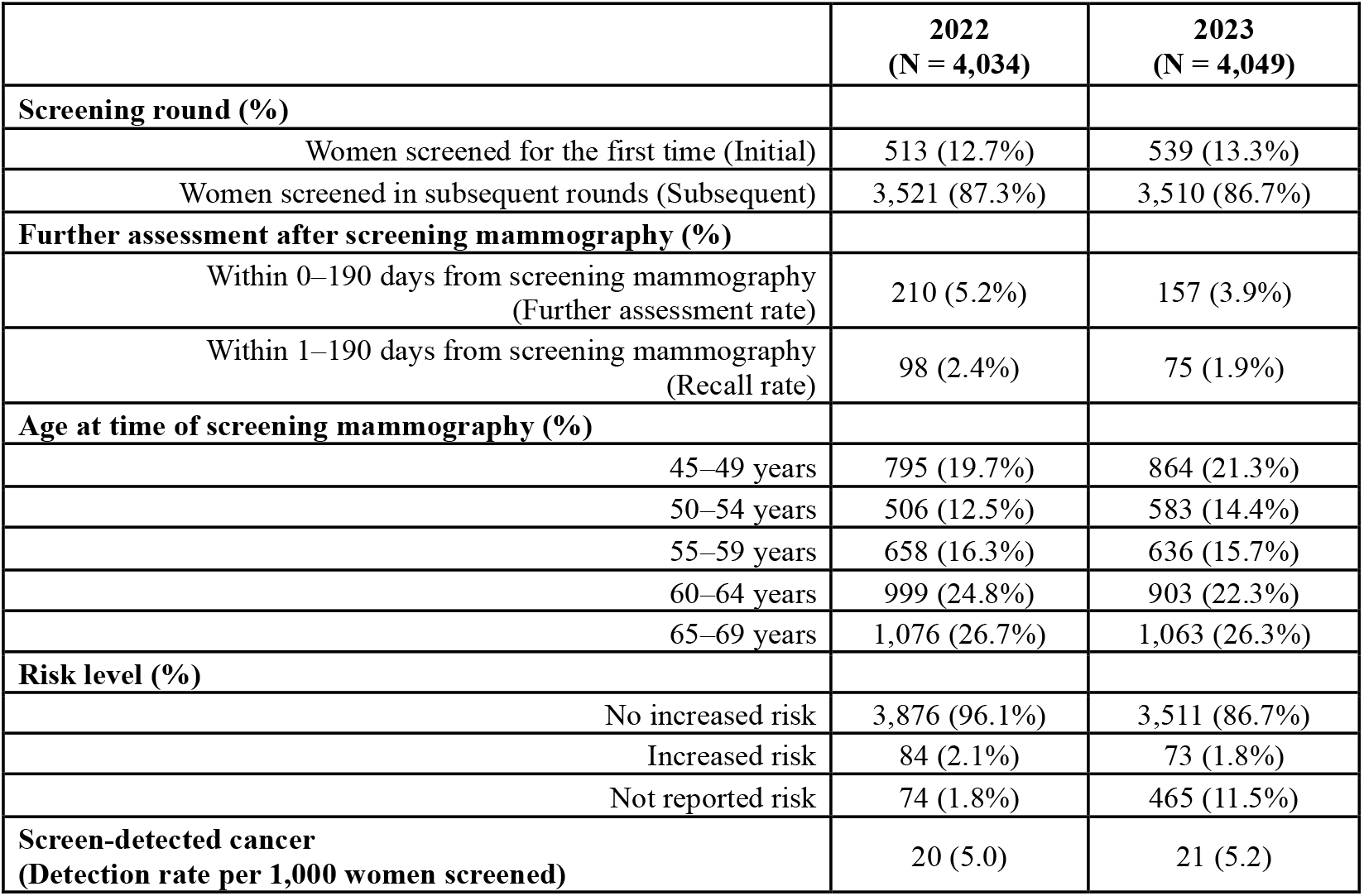
Basic characteristics of the women aged between 45 and 69 years.

|  | **2022**<br>**(N = 4,034)** | **2023**<br>**(N = 4,049)** |
| --- | --- | --- |
| **Screening round (%)** |  |  |
| Women screened for the first time (Initial) | 513 (12.7%) | 539 (13.3%) |
| Women screened in subsequent rounds (Subsequent) | 3,521 (87.3%) | 3,510 (86.7%) |
| **Further assessment after screening mammography (%)** |  |  |
| Within 0–190 days from screening mammography<br>(Further assessment rate) | 210 (5.2%) | 157 (3.9%) |
| Within 1–190 days from screening mammography<br>(Recall rate) | 98 (2.4%) | 75 (1.9%) |
| **Age at time of screening mammography (%)** |  |  |
| 45–49 years | 795 (19.7%) | 864 (21.3%) |
| 50–54 years | 506 (12.5%) | 583 (14.4%) |
| 55–59 years | 658 (16.3%) | 636 (15.7%) |
| 60–64 years | 999 (24.8%) | 903 (22.3%) |
| 65–69 years | 1,076 (26.7%) | 1,063 (26.3%) |
| **Risk level (%)** |  |  |
| No increased risk | 3,876 (96.1%) | 3,511 (86.7%) |
| Increased risk | 84 (2.1%) | 73 (1.8%) |
| Not reported risk | 74 (1.8%) | 465 (11.5%) |
| **Screen-detected cancer**<br>**(Detection rate per 1,000 women screened)** | 20 (5.0) | 21 (5.2) |

In 2023 (AI-assisted assessment), the cancer detection rate increased slightly to 5.2 breast cancers detected per 1,000 women screened, up from 5.0 in 2022 (non-AI assessment). This increase is not statistically significant, even when adjusting for the effects of the women’s age, risk level, and screening round (*p* = 0.919; OR (95% CI): 1.034 (0.542–1.974)) (Figure 2).

**Figure 2:**
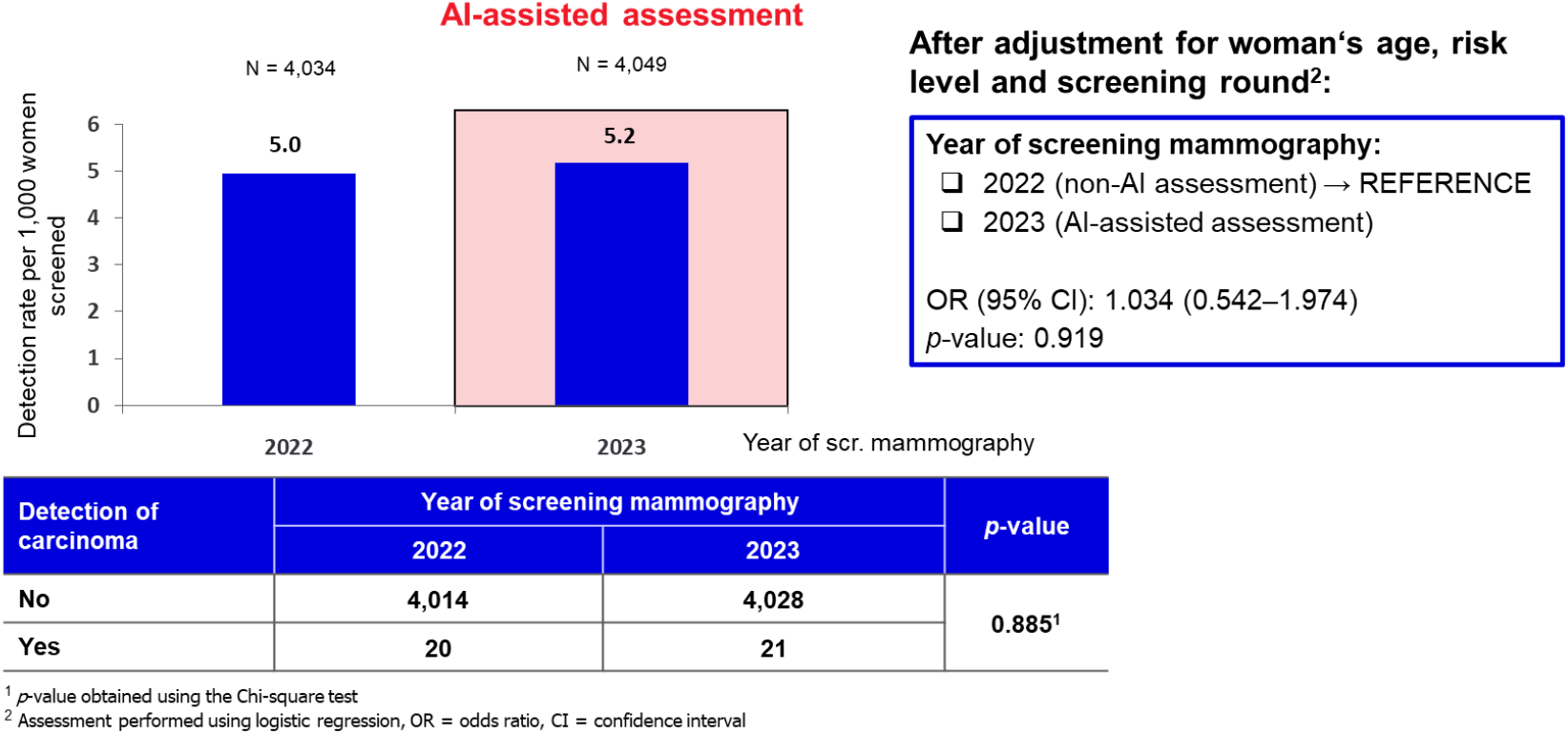
Comparison of breast cancer detection rates (CDR) with and without AI assistance.

The proportion of additional examinations (FAR) in 2023 decreased compared to 2022. In 2023 (AI-assisted assessment), the proportion of supplementary examinations declined to 3.9% from 5.2% in 2022 (non-AI assessment). This difference between the proportions of additional examinations in 2022 (non-AI assessment) and 2023 (AI-assisted assessment) is statistically significant, even when adjusting for the effects of the women’s age, risk level, and round of the screening (*p* < 0.001; OR (95% CI): 0.665 (0.529–0.836)) (Figure 3).

**Figure 3:**
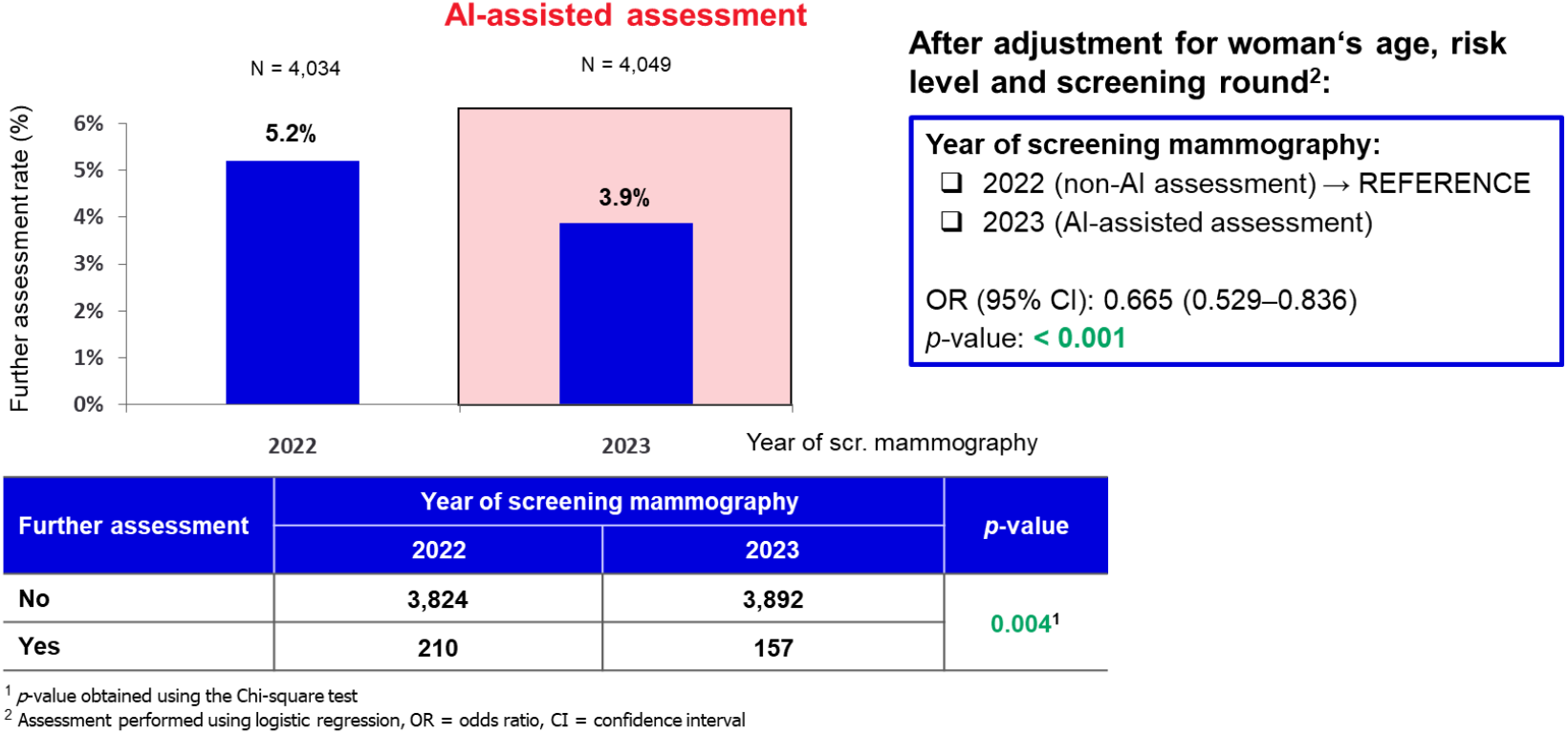
Comparison of further assessment rates (FAR) with and without AI assistance.

There was also a non-significant decrease in the proportion of women invited for a follow-up examination (RR) in 2023 (AI-assisted assessment) to 1.9% from 2.4% in 2022 (non-AI assessment). The difference between the proportions of women invited for a follow-up examination in 2022 (non-AI assessment) and 2023 (AI-assisted assessment) is borderline statistically significant, even when adjusting for the effects of the woman’s age, risk level, and round of the screening (*p* = 0.083; OR (95% CI: 0.754 (0.548–1.037)) (Figure 4).

**Figure 4:**
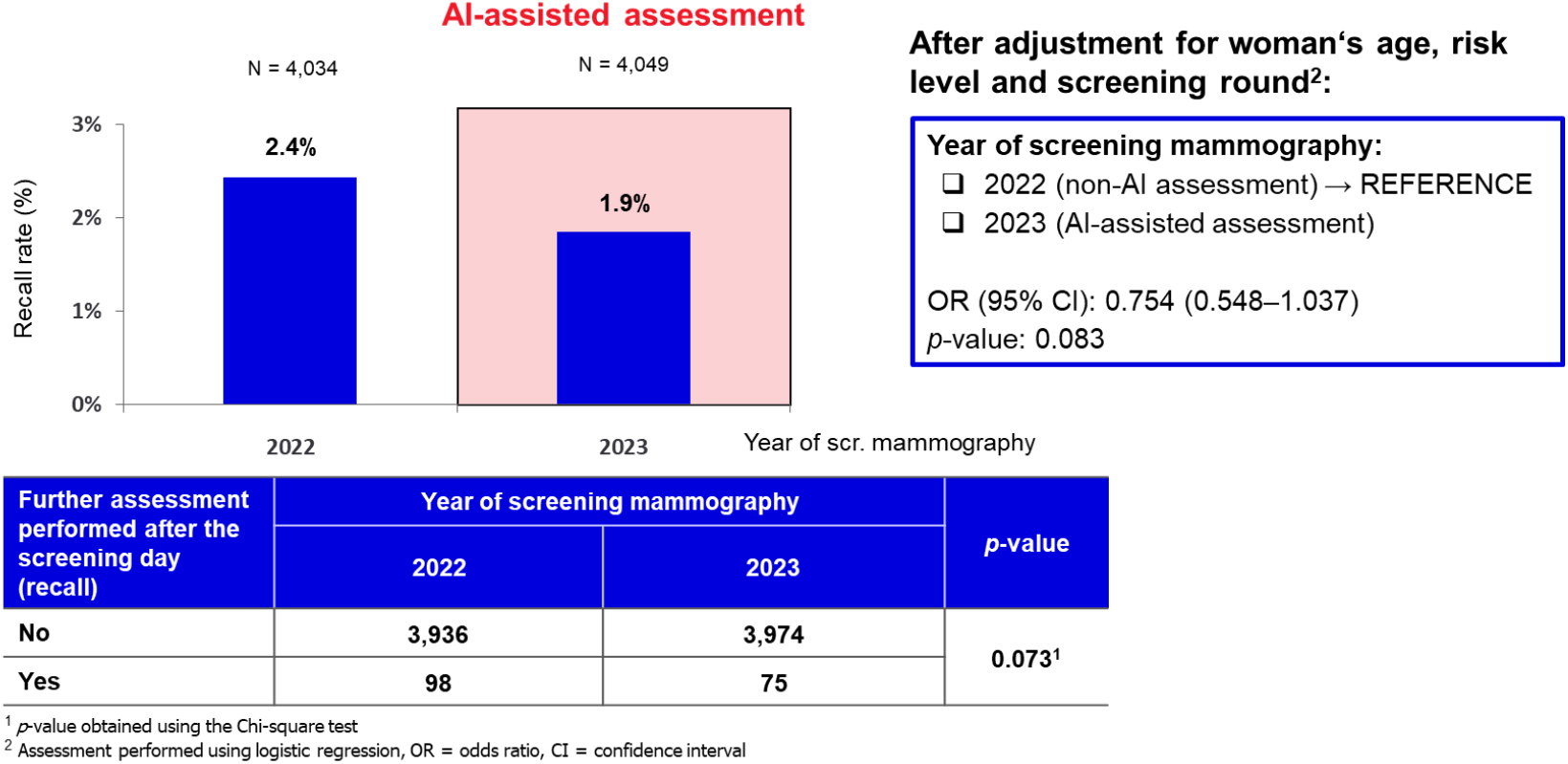
Comparison of recall rates (RR) with and without AI assistance.

## Discussion

The results of our study suggest that although stand-alone AI software could have a high false positivity rate, for example, marking some stationary lesions as high-risk, its use in routine clinical practice does not lead to an increase in the number of additional examinations. The values that indicate the number of additional tests such as RR and FAR in our study did not increase; on the contrary, they decreased, while the cancer detection rate was maintained. It was not possible to obtain information about high risk from a certain number of women. Therefore, logistic regression analysis without the women’s risk level predictor was performed; however, the results remained unchanged and, given the importance of this variable as an explanatory factor, it was retained in the final model. The gold standard in most European countries is the reading of screening mammography by two experienced radiologists [6]. The primary goals of this approach are to reduce the number of unnecessary additional examinations, maximise the detection of early invasive breast cancer, and minimise the incidence of interval breast cancer (false negative cases). One study using the iCAD system confirmed that the sensitivity of this system for the evaluation of mammography was comparable to human reading but its specificity was lower. Combining human reading with AI increased sensitivity compared with reading without AI support [8]. In our study, double reading by a radiologist with AI as an additional reader was evaluated.

Our results are in line with the results of most published studies [9-16]. In many studies similar results for the recall rate or the number of additional examinations after screening mammography were obtained. Some studies reported a reduction in the number of additional examinations [11,12], while others reported no impact when using AI software [14,15], one study reported a low increase in RR [13]. Regarding the CDR value, most studies reported an increase in CDR when using an AI-based system [9-11,13-15].

In an observational multicentre implementation study (PRAIM), AI-supported double reading was associated with a higher CDR without a negative impact on RR [12]. The radiologists in the AI-supported screening group achieved a breast cancer detection rate of 6.7 per 1,000, which was higher than the rate (5.7 per 1,000) achieved in the control group. The RR in the AI group was 37.4 per 1,000, which was lower than the rate (38.3 per 1,000) in the control group. It strongly indicates the ability of AI to exert a positive influence on mammographic screening metrics. Similar results are also reported in our study.

Some studies, MASAI study and the others, report a reduction in workload when using AI, particularly when using tomosynthesis [10,11,15,17]. This was not considered in our study.

The lower FAR and RR in the AI-using group in our study represent key findings. It is worth considering whether this could lead to a reduction in the number of unnecessary additional examinations and biopsies. Conversely, increased sensitivity when using AI in screening could lead to overdiagnosis [18].

On the other hand, the positive impact of mammography-based AI for non-invasive prediction of the breast cancer subtype was reported, opening up the way for a promising alternative to biopsies and for personalised treatment plans [19]. It is exciting to imagine a future where AI is used successfully to avoid unnecessary biopsies in thousands of women [18,19]. This is a very bold statement, but several publications in other fields of radiology have discussed the use of radiomics, as well as mentioning the idea of using radiomics in combination with AI in the future to develop approaches resembling virtual biopsies to avoid unnecessary examinations or biopsies [20,21].

In the light of the growing body of evidence, the Guideline Development Group (GDG) of the European Commission Initiative on Breast Cancer (ECIBC) still favours the use of AI-supported double readings (with consensus or arbitration for non-consensus readings) over non-AI-supported double readings or AI-supported single readings by a radiologist [16].

The strength of our study is that it is a study in an organised screening system that includes consecutive women. On the other hand, our study also has some limitations. It is a retrospective study, which may be burdened by biases such as confounding by unmeasured factors. It is a study in one specific centre. With regard to the general applicability of the results, it must be mentioned that this is a centre with a large number of surgical procedures for breast cancer. The radiologists at this centre read a large number of diagnostic mammograms with carcinomas. On the other hand, the number of screening mammograms per year is lower. This centre has two radiologists with more than ten years of experience, but also has an equal number of radiologists with less than ten years of experience. The further assessment in Czech Republic is rather high compared with other European screening programmes [22], which could have led to a more pronounced reduction of further assessment rates than observed in previous studies.

As noted above, AI-based systems have the potential to improve screening results, reduce the number of false negatives and positives, and detect subtle abnormalities that are missed by human observers. Given the evidence from numerous retrospective studies, it is likely that artificial intelligence could play a central role in the future of breast cancer screening programmes, for example by replacing one of two human readers. The false positivity of these systems has no impact on quality indicators in real clinical practice, if these systems are used as a reading assistant, according to our retrospective study and also according to published studies.

In the future, a prospective randomised study at the same screening centre is planned, also for determining the histopathological subtypes of the cancers that are detected, to evaluate the above-mentioned overdiagnosis.

## Conclusion

As an upcoming future technology, AI has the potential to improve the breast cancer diagnostic process by increasing accuracy and reducing extra examinations and radiologists’ workload. This advance is emerging as a response to the increasing demand for effective screening methods amid challenges such as shortages of radiologists and a growing patient population. Adding an AI system as a support for a standard double reading could improve mammography screening metrics.

Despite the technological advances, circumspection and caution about implementing these results into real clinical practice are necessary.

## Data Availability

All anonymized data and scripts are publicly available at Zenodo in accordance with Open Science principles at: https://doi.org/10.5281/zenodo.20327197

## Abbreviations

2D FFDM: 2D Full-Field Digital Mammography
AI: artificial intelligence
AI-CAD: AI-based computer-aided detection
AIR: abnormal interpretation rate
CDR: cancer detection rate
CI: confidence interval
DBT: digital breast tomosynthesis
ECIBC: The European Commission Initiative on Breast Cancer
FAR: further assessment rate
GDG: Guidelines Development Group
IBA LF: Institute of Biostatistics and Analyses, Faculty of Medicine
OR: odds ratio
PPV: positive predictive value
RR: recall rate

## Acknowledgement

Not applicable.

## Ethics approval and consent to participate

This retrospective study was approved by the institutional Ethics Committee of the University Hospital and Faculty of Medicine and Dentistry of Palacký University Olomouc (reference no. 52/26). All procedures were performed in accordance with relevant guidelines and regulations.

The patients provided written informed consent to the examination, including consent to using documentation anonymously for scientific and statistical purposes.

Some of the authors have significant statistical expertise.

## Declaration of the Use of Artificial Intelligence

In the preparation of this work, the tool ChatGPT (model GPT-5, OpenAI, 2025) was used. The model was utilized for grammar revision and stylistic editing of the text. Following the use of this tool, the author reviewed the content and assumes full responsibility for it.

